# Psychological Stress-Associated Ceramide and Diacylglyceride Lipotoxicity as Contributors to First Episode Depression Pathophysiology: A neuroimmune-Metabolic-Oxidative Stress (NIMETOX) Perspective

**DOI:** 10.64898/2026.05.18.26353450

**Authors:** Visuthsiri Sirivatanapa, Pannipa Janta, Asara Vasupanrajit, Chavit Tunvirachaisakul, Sira Sriswasdi, Rossarin Tansawat, Andre F Carvalho, Yingqian Zhang, Michael Maes

**Affiliations:** Department of Psychiatry, Faculty of Medicine, Chulalongkorn University, Bangkok, Thailand; Ph.D. Program in Mental Health, Department of Psychiatry, Faculty of Medicine, Chulalongkorn University, Bangkok, Thailand; Center of Excellence in Metabolomics for Life Sciences, Chulalongkorn University, Bangkok, 10330, Thailand; Department of Food and Pharmaceutical Chemistry, Faculty of Science, Chulalongkorn University, Bangkok, Thailand; Cognitive Impairment and Dementia Research Unit, Faculty of Medicine, Chulalongkorn University, Bangkok, Thailand; Center of Excellence in Computational Molecular Biology, Division of Research Affairs, Faculty of Medicine, Chulalongkorn University, Bangkok, Thailand; Center for Artificial Intelligence in Medicine, Division of Research Affairs, Faculty of Medicine, Chulalongkorn University, Bangkok, Thailand; Innovation in Mental and Physical Health and Clinical Treatment (IMPACT) Strategic Research Centre, School of Medicine, Barwon Health, Deakin University, Geelong, VIC 3220, Australia; International NIMETOX Center, Sichuan Provincial Center for Mental Health, Sichuan Provincial People’s Hospital, School of Medicine, University of Electronic Science and Technology of China, Chengdu 610072, China; Key Laboratory of Psychosomatic Medicine, Chinese Academy of Medical Sciences, Chengdu, 610072, China; Department of Psychiatry, Medical University of Plovdiv, Plovdiv, Bulgaria; Research Institute, Medical University of Plovdiv, Plovdiv, Bulgaria; Research and Innovation Program for the Development of MU – PLOVDIV (SRIPD-MUP), Creation of a network of research higher schools, National Plan for Recovery and Sustainability, European Union – NextGenerationEU, Medical University of Plovdiv, Plovdiv, Bulgaria, Europe; Kyung Hee University, Seoul, Dongdaemun-gu, South Korea

**Keywords:** major depressive disorder, neuroimmune, antioxidants, oxidative and nitrosative stress, inflammation, biomarkers

## Abstract

**Background:** Aberrations in neuro-immune, metabolic, and oxidative stress (NIMETOX) pathways are implicated in major depressive disorder (MDD). First-episode simple dysmood disorder (FE-SDMD) without metabolic syndrome offers a unique model to investigate early lipid alterations underlying NIMETOX pathophysiology.

**Methods:** Plasma samples were collected from 88 university students (44 FE-SDMD, 44 healthy controls). Participants underwent comprehensive psychiatric and psychological assessments, including adverse childhood experiences (ACEs), negative life events (NLEs), depression, anxiety, suicidal behaviors, and insomnia. Untargeted lipid profiling was performed using LC-QTOF-MS, while indices of oxidative and nitrosative stress (ONS) and lecithin–cholesterol acyltransferase (LCAT) activity were assessed. Data was analyzed using machine learning approaches with recursive feature elimination and cross-validation.

**Results:** FE-SDMD was characterized by increased ceramides (CER), diacylglycerides (DAG), triacylglycerides (TG), sphingomyelins (SM), bis-monoacylglycerol phosphates (BMP), cholestone, and fatty-acyl amino acids (FAAA). DAG, CER, and BMP were the strongest predictors of depression severity and physiosomatic symptoms, whereas cholestone, CER, and SM predicted suicidal behaviors. These lipid modules, together with lowered LCAT and increased ONS, explained substantial variance in depression severity (46.4%), physiosomatic symptoms (42.4%), cognitive-affective symptoms (37.9%), suicidal behaviors (30.1%), insomnia (32%), and anxiety (19.5%). ACEs and NLEs were strongly associated with CER (p<0.001), DAG (p<0.01), and cholestone (p<0.01).

**Conclusion:** Early-stage MDD is characterized by distinct lipid dysregulations linked to psychosocial stress exposure, oxidative and nitrosative stress, and an indicant of impaired reverse cholesterol transport. These lipid modules may serve as early biomarkers and therapeutic targets in vulnerable populations.

## Introduction

Major depressive disorder (MDD) is a complex and heterogeneous condition characterized by disturbances in neuroimmune, metabolic, and oxidative stress pathways, collectively termed neuroimmune–metabolic–oxidative stress (NIMETOX) pathways (Maes, Almulla, You, et al., 2025). Recent evidence suggests that MDD may be divided into two subtypes: simple dysmood disorder (SDMD), characterized by milder symptoms and fewer recurrent episodes, and major dysmood disorder (MDMD), which is associated with more severe symptoms and frequent recurrences (Maes et al., 2023). These subtypes not only differ clinically but also show distinct immune-metabolic profiles (Maes et al., 2022). MDMD is accompanied by increased activation and sensitization of the immune-inflammatory response system (IRS), whereas SDMD and first-episode SDMD (FE-SDMD) are mainly characterized by reduced activity of the compensatory immune-regulatory system (CIRS) (Maes et al., 2022; Maes, Vasupanrajit, et al., 2025). In addition, patients with FE-SDMD show increased oxidative and nitrosative stress (ONS), as indicated by elevated malondialdehyde (MDA), nitric oxide metabolites (NOx), and reduced antioxidant defenses measured by total antioxidant potential (TRAP) (Brinholi et al., 2024).

Research has shown that lipid abnormalities occur in individuals with MDD, including reduced lecithin–cholesterol acyltransferase (LCAT) activity (Maes et al., 1994; Maes, Vasupanrajit, et al., 2024). LCAT, activated by apolipoprotein A1 (ApoA1), esterifies free cholesterol on high-density lipoprotein (HDL) particles, thereby promoting HDL maturation and the formation of HDL cholesterol (HDLc) (Zannis et al., 2006). Mature HDL particles can bind paraoxonase 1 (PON1), an antioxidant enzyme that protects lipids from oxidative damage (Gugliucci & Menini, 2015; Jaouad et al., 2003). These processes support reverse cholesterol transport (RCT), a key pathway involved in cholesterol removal and protection against oxidative stress and inflammation (Almulla et al., 2023). PON1, HDL-cholesterol, LCAT and ApoA1 are all decreased in MDD, suggesting impairments in RCT (Almulla et al., 2023; Maes, Zhou, et al., 2024; Moreira et al., 2019). Moreover, adverse childhood experiences (ACEs) are associated with lower HDLc levels, weakened antioxidant defenses, and increased atherogenicity, suggesting that early-life stress may contribute to persistent lipid and oxidative abnormalities later in life (Maes, Jirakran, et al., 2024; Moraes et al., 2018).

Lipidomics, metabolomics, and fatty acid (FA) profiling provide a comprehensive view of fatty acid-lipid disturbances in MDD. Previous studies show depletion of structural membrane lipids, omega-3/omega-6 substrates, PUFA remodeling pools, very long chain FAs, and odd-chain FAs in MDD (Maes, Niu, Almulla, et al., 2026). These FA modules were negatively associated with increased ceramide (CER) and diacylglyceride (DAG) levels and phospholipid (PL) remodeling but positively with lowered (acyl)carnitines and plasmalogens (Maes, Niu, Almulla, et al., 2026). These changes were independent of metabolic syndrome (MetS) or body mass index (BMI). A previous non-targeted lipidomics study found that individuals with MDD showed increased levels of DAG, triglycerides (TG), and phosphatidylcholine (PC), with decreased sphingomyelins (SM) (Liu et al., 2016). Importantly, a composite FA-lipotoxicity index—combining elevated lipotoxicity and CER with reduced plasmalogens (reflecting diminished antioxidant defenses) and depleted structural fatty acids—was strongly associated with IRs activation in MDD (Maes, Niu, Almulla, et al., 2026). Given that IRS processes can trigger downstream CER toxicity (Bernal-Vega et al., 2023; Zorkina et al., 2024), these lipids may represent a critical part within the NIMETOX framework.

Altered lipid metabolism also appears even in sub-clinical stages or in first episode (FE) MDD. Individuals diagnosed with MDD showed higher erythrocyte omega-6 and omega-9 fatty acids including arachidonic, dihomo-γ-linolenic, elaidic, and oleic acids (L. Wang et al., 2022). Moreover, Maes, Vasupanrajit, et al. (2024) showed that FE-SDMD students exhibited increased TG, free cholesterol, and atherogenicity index, accompanied by a reduced LCAT index (Maes, Vasupanrajit, et al., 2024). Moreover, increased atherogenicity and lowered LCAT activity were positively linked with overall depression severity and/or suicidal behaviors (Maes, Vasupanrajit, et al., 2024). Dysregulation of lipid pathways was less pronounced in those with mild MDD relative to those with severe MDD (Liu et al., 2016). Taken together, these findings suggest that lipid dysregulation may precede the onset of full blown MDD and that degrees of dysregulation may differ across symptom burden and disease staging. This makes lipid profiling a potential window into the early pathogenesis.

However, comprehensive studies of lipid profiles in individuals with FE-SDMD remain limited. The investigation of FE-SDMD without MetS may provide unique clinical and biological insights, since these individuals are free from the confounding effects of long-term medication, recurrence of illness (ROI), and comorbid metabolic aberrations. This provides an opportunity to identify key lipid alterations that may drive the early pathophysiology of MDD. Moreover, FE-SDMD is associated with distinct cytokine, chemokine, and growth factor profiles compared with MDMD and healthy controls, further highlighting unique immune–metabolic signatures in this early-stage subtype (Maes, Vasupanrajit, et al., 2025).

Thus, the objective of the current research is to characterize the plasma lipidomics profile of FE-SDMD individuals relative to healthy controls using liquid chromatography quadrupole time-of-flight mass spectrometry (LC-QTOF-MS) analysis. Given the established role of lipids including free cholesterol and TG and diminished RCT, antioxidant defenses and plasmalogen in MDD (Maes, Niu, Almulla, et al., 2026), we hypothesize that FE-SDMD could be accompanied by increased CER, SM, DAG, and signaling lipids. Furthermore, since CER and DAG are bioactive signaling lipids, we hypothesize that their concentrations could be positively associated with the key phenome features of MDD, i.e., overall severity of illness, pure cognitive-affective symptoms, physiosomatic symptoms, suicidal behaviors, insomnia, anxiety, and possibly with psychological stressors, including ACEs and negative life events (NLEs).

## Materials and Methods

Plasma samples and clinical data used in this analysis were originally collected as part of a completed study (Vasupanrajit et al., 2023) which was overseen by the Institutional Review Board (IRB) of the Faculty of Medicine, Chulalongkorn University (IRB No. 351/63). Participant recruitment occurred between November 2021 and February 2023. After study closure, de-identified residual samples along with their associated clinical and biomarker data were made available for the current research. The current research was reviewed and approved by the Board (IRB) of the Faculty of Medicine, Chulalongkorn University for use of these materials (IRB No. 045/68).

### Participants

Participants were 88 Thai-speaking students enrolled at various faculties of Chulalongkorn University. The students were both males and females aged between 18 and 35 years. Students with FE-SDMD (n=44) were outpatients of the Department of Psychiatry, King Chulalongkorn Memorial Hospital, Bangkok, Thailand. We recruited 123 consecutively admitted MDD outpatients and of those 35 were excluded because they already suffered previously from an MDD episode, MDMD or MetS. Healthy controls (HC) were 44 university students. Exclusion criteria were participants with metabolic syndrome, current breastfeeding or pregnancy, endocrine or autoimmune and immune disorders, neurological disorder, cardiovascular disorder, signs of severe suicidal ideation, or other major psychiatric disorder.

### Data Collection and Clinical Assessments

The Mini International Neuropsychiatric Interview (M.I.N.I.) (Kittirattanapaiboon, 2004) was used to diagnose axis 1 DSM-IV-TR disorders (depressive disorders, anxiety disorders, obsessive-compulsive and related disorders, trauma and stress related disorders, eating disorders, and substance use disorders). Depression and anxiety were diagnosed with the DSM-5 (American Psychiatric Association, 2022).

State-Trait Anxiety Inventory (STAI) (Spielberger et al., 1971) was used to evaluate anxiety severity in all participants, while Hamilton Depression Scale (HAM-D) (Hamilton, 1960) and Beck depression inventory-II (BDI-II) (Beck, 1996) were used to assess depression severity. The sum of the BDI-II items, except irritability, appetite, tiredness, interest in sex, and sleep, was used to construct a “pure BDI” or “restricted cognitive-affective” score, which reflects the cognitive depressive symptoms. The physiosomatic symptoms were a principal component score constructed using 3 HAM-D items (somatic anxiety, gastro-intestinal somatic, and general somatic) and 2 BDI-II items (loss of energy and fatigue). The Insomnia Severity Index (ISI) (Morin et al., 2011) is a 7-item scale that measures insomnia symptoms and severity which offers Cronbach alpha from 0.90 to 0.91. The Thai translation was provided by Mapi Language Services (2015). The Columbia-Suicide Severity Rating Scale (CSSRS) (Posner et al., 2011) was used to assess suicidal ideation (SI) and attempts (SA) within the past 1 month and lifetime. The Thai version was provided by The Columbia Lighthouse Project (2016). We constructed suicide behavior (SB) principal components as described previously (Vasupanrajit et al., 2023): the 10-item CSSR SI lifetime was used to construct PC SI Lifetime; 10-item CSSR SI in the past 1 month to construct PC SI current; CSSR SA lifetime item 1,4,6,8,9 to construct PC SA lifetime; CSSR SA in the past 1 month item 1,2,4,6,8,9 to construct PC SA current (Vasupanrajit et al., 2023). PC SB lifetime was calculated by PC SI Lifetime + PC SA Lifetime, while PC current SB was computed by the formula PC SI current + PC SA current. Finally, total suicide behavior was calculated by PC SB lifetime + PC SB current (Vasupanrajit et al., 2023). Overall severity of depression (OSOD) is a comprehensive phenome index constructed by extracting the first PC from the total HAM-D, BDI-II, STAI, ISI, and current SB scores (Vasupanrajit et al., 2023).

The Adverse Childhood Experiences (ACE) Questionnaire was used to assess different ACE domains, namely, child abuse (emotional, physical and sexual abuse), and neglect (emotional and physical neglect) (Felitti et al., 1998). The questionnaire was translated into the Thai language by Rungmueanporn et al. (2019) and showed an internal consistency reliability of 0.79 for abuse domain, and 0.82 for the neglect domain. We used the sum of these 5 ACE domains as an index of ACE load (Maes, Almulla, et al., 2024). The Thai translation (Boonyamalik, 2005) of the Negative Event Scale (Maybery, 2003) was used to measure interpersonal and non-interpersonal negative life events (NLEs). The six interpersonal NLEs were problems with friends, boy/girlfriend, lecturers, parents, other students, and relatives. The four non-interpersonal NLEs were money, course, course interest, health, and academic limitations. Then, PC NLEs self (money + health), PC relationship (friends + parents + other students + relatives), and PC NLEs academic (course + course interest + academic limitations) were calculated.

The Joint Scientific Statement of the American Heart Association and the National Heart, Lung, and Blood Institute criteria were used to diagnose and assess MetS, a condition characterized by a cluster of cardiovascular and metabolic risk factors, including abdominal obesity, dyslipidemia, hypertension, and insulin resistance (Alberti et al., 2009). MetS is diagnosed when three or more of the following are present: abdominal obesity (waist circumference >90 cm in men, >80 cm in women), triglycerides ≥150 mg/dL, HDL <40 mg/dL (men), <50 mg/dL (women), blood pressure ≥130/85 mmHg, and fasting glucose ≥100 mg/dL. MetS was used as an exclusion criterion.

### Assays

Following an overnight fast of 8 hours, blood samples were collected at 7.30 a.m. and kept at –80. Detailed data collection and protocol of this study can be found in our previous work (Vasupanrajit et al., 2023). MDA, NOx, TRAP, total and free cholesterol were measured as described previously (Brinholi et al., 2024). The ONS index is a composite score calculated by z score of MDA (z MDA) + z NOx – z TRAP (Brinholi et al., 2024). LCAT index which reflects the esterified cholesterol ratio was calculated by (1 – free cholesterol / total cholesterol) × 100 (Maes et al., 1994).

### Lipid Profiling Using LC-QTOF-MS

#### Chemicals and Reagents

Methanol and water were obtained from Honeywell (USA). Ethanol, acetonitrile, and butylated hydroxytoluene (BHT; Pharmaceutical Secondary Standard, Certified Reference Material) was obtained from Supelco (Germany). Ammonium acetate was purchased from Merck (Germany). The internal standard mixture, UltimateSPLASH™ ONE, was purchased from Avanti Polar Lipids (USA).

#### Sample Preparation and Lipid Extraction

Lipid extraction was performed using a modified butanol:methanol method adapted from (Perera et al., 2023). A 50 mL working extraction solvent was prepared by mixing butanol and methanol (1:1). After vortex, 2 mL of the solvent was replaced with 2 mL of 10 mM ammonium acetate. Next, internal standard solution (Ultimate Splash standards) was added. After vortex, 0.01% BHT (dissolved in acetonitrile) was added to the mixture.

Plasma samples were thawed at 4□, and 20 μL of each sample was transferred to an Eppendorf tube. A pooled biological quality control (PBQC) sample was created by combining 20 µL aliquots from all study samples. Lipid extraction was initiated by adding 180 μL of the previously prepared extraction cocktail to each Eppendorf tube, followed by vortex, shaking, and centrifugation. The supernatants were transferred to glass vials with inserts for LC-MS/MS analysis. A blank sample was inserted and analyzed at the beginning of each sequence. PBQC was inserted into the analysis sequence every ten real samples.

Lipid profiling was achieved using a Shimadzu LC-40D-X3 system (Shimadzu, Japan) coupled to a ZenoTOF 7600 mass spectrometer (SCIEX, Concord, CA). The TOF system was equipped with OptiFlow turbo ion source using electrospray ionization (ESI) mode. Chromatographic separation was performed on a Kinetex C18 column (100 × 2.1 mm, 1.7 µm; Phenomenex, Australia) maintained at 40°C. The mobile phases of (A) acetonitrile:water (60:40) with 10 mM ammonium acetate and (B) consisted of isopropanol:acetonitrile (90:10) with 10 mM ammonium acetate were used. The flow rate was set to 0.4 mL/min with injection volume of 5 μL. Parameters of ESI were as follows: source temperature = 350°C, curtain gas = 30 psi, collision gas = 7 psi, ion source gas 1 = 45 psi, and ion source gas 2 = 40 psi.

Full scan TOFMS data were acquired in positive ion mode (m/z 50–1500) using ion spray voltage of 5500 V, accumulation time 0.05 s, declustering potential of 80 V with 10 V spread, collision energy of 12 V. TOFMS/MS acquisition was performed in positive ion mode using electron-based fragmentation (EAD). The source parameters were set as follows: spray voltage 5500 V, mass range 80–1200 m/z, accumulation time 0.035 s, and a declustering potential of 80 V with 10 V spread. The EAD settings were an electron beam current of 7500 nA, electro kinetic energy of 15 eV, and an electron transfer control of 100. The EAD radio frequency was 80 Da with a reaction time of 30 ms. The Zeno trap threshold was set to 80,000 counts per second.

#### Data Preprocessing

Peak detection, spectral deconvolution, data alignment, and lipid annotation of the raw LC-MS data were carried out using MS-DIAL version 5.50. For each annotated lipid, the area under the curve (AUC) was subjected to 50% filtering, zero imputation using 1e-9, centered-log-ratio (CLR) transformation, and z-standardization prior to downstream statistical analyses, including regression analyses.

### Statistics

Categorical variables were analyzed using contingency tables with Chi-square tests. Continuous variables, including lipidomics data, were compared using Wilcoxon tests or one-way analysis of variance (ANOVA) as appropriate. Associations between continuous variables were assessed using Pearson correlation coefficients. To control multiple testing, p-values were adjusted using the False Discovery Rate (FDR) method. Partial least squares discriminant analysis (PLS-DA) was applied to assess multivariate separation between FE-SDMD and control groups based on the lipidomics data. Variable Importance in Projection (VIP) scores were calculated to identify lipids contributing most to group discrimination, and contribution scores were derived to quantify the influence of each lipid for all cases separately. Model robustness and potential overfitting were evaluated using permutation testing (n = 1000), comparing observed R² and Q² values to distributions obtained from randomly permuted class labels. VIP scores >1 were considered significant contributors. PLS-regression was performed to model associations between lipid modules and key symptom domains, including OSOD, pure BDI, physiosomatic symptoms, and suicidal behaviors. Lipids with high VIP in the PLS regression were interpreted as key predictors of these key domains. Random forest (RF) models were implemented to classify FE-SDMD versus control participants. Recursive feature elimination (RFE) with Cross-Validation (RFECV) was applied to iteratively select the most relevant variables while minimizing overfitting. Model performance was assessed using 5-fold cross-validation, and variable importance measures were computed to rank lipids according to their contribution to classification accuracy. To reduce the risk of overfitting, the dataset was partitioned into training and testing subsets using a 70:30 split. Balanced accuracy, sensitivity, specificity, and area under the receiver operating characteristic curve (AUC) were reported to evaluate model performance.

To derive the lipid modules that significantly predict the key phenome features, we applied both manual multiple regression and automated regression techniques. Ridge regression was conducted with λ = 0.1 and a tolerance of 0.4. In addition, we carried out forward-stepwise automatic linear modeling, using an overfit criterion for determining variable entry and removal and setting the maximum number of effects to 5. To further limit overfitting, we implemented a “best subset” procedure based on 8 lipidomic modules obtained from the SPSS regression. Following these procedures, we performed a manual regression analysis to evaluate model statistics, including the F statistics, degrees of freedom, p-value, and R^2^ explained variance. We also inspected each predictor’s standardized beta coefficient, t-statistics, and exact p-value. Multivariate normality of the final models was examined using residual distributions and P–P plots. To identify collinearity, we reviewed the variance inflation factor and tolerance values. Heteroskedasticity was evaluated using the White test and the modified Breusch–Pagan test. The significance level for all statistical tests was set at 0.05 (two-tailed). Statistical analyses were conducted using IBM SPSS Statistics version 30, R (version 4.0), Statistica version 14, the scikit-learn library in Python, and the Unscrambler. Visualization techniques comprised the R packages heatmap/pheatmap for cluster heatmaps, while PLS-DA figures were produced using OmicStudio (https://www.omicstudio.cn/tool). Heatmaps and bar plots were created using Seaborn and Matplotlib (Python).

## Results

### Demographic and clinical data

The Electronic Supplementary File (ESF) Table 1 shows the clinical, demographic and biomarker data of the FE-SDMD patients in the current study. There were no significant differences in sex ratio, age, BMI, education, MetS, and smoking status between FE-SDMD patients and controls. Patients with FE-SDMD had elevated OSOD, physiosomatic symptoms, pure BDI, ISI, suicidal behavior and STAI scores as compared with controls. The ONS index was significantly higher in FE-SDMD patients than controls, whereas the LCAT index was significantly decreased.

**Table 1.** Construction of eight functional lipid modules.

| Modules | Members (synonym) | Functions |
| --- | --- | --- |
| Diacylglycerides (DAG) | DG 56:2<br>DG 62:3<br>DG O-35 | DAG are potent signaling lipids because they can activate protein kinase C (PKC), which is a central switch to insulin resistance, endoplasmic reticulum (ER) stress, and inflammation. DAG are precursors to TG and phospholipid resynthesis. DAG are harmful lipid signaling molecules that disrupt lipid buffering and can act together with CER to exacerbate neuroimmune and metabolic stress. |
| Ceramides (CER) | CER 38:0<br>CER 50:4<br>CER 56:10<br>CER 65:6<br>CER 46:8<br>CER 72:5<br>CER 60:5<br>CER 65:7 | CER is membrane lipid. In excess, CER are lipotoxic molecules that rigidify membranes, disrupt insulin receptor signaling, permeabilize mitochondria through CER channels formation, and block beta-oxidation. Excess CER drive cells from adaptive lipid remodeling into inflammatory, apoptotic, and metabolically dysfunctional states. CER can be converted to SM and vice versa. |
| Bismonoacylglycerol phosphates<br>(glycerophospholipids; BMP) | BMP 27:2 | BMP is abundant in lysosomal membrane's intraluminal vesicles, where it supports the sorting and breakdown of lipids, including sphingolipids and cholesterol. BMP contributes to the regulation of CER metabolism, sphingolipid turnover, and lysosomal cholesterol efflux. Because of this, elevated BMP levels may reflect an increased influx of these lipids into lysosomes. |
| Fatty-acyl amino-acid conjugates (FAAA) | NaGlySer 53:6 | FAAAs like N-acyl glycine-serine conjugate (NaGlySer) are signaling lipids associated with anti-inflammation and detoxification. In response to dysfunctional beta-oxidation, FAAA often increases to reduce excess polyunsaturated fatty acids influx. When co-occur with increased CER, decrease plasmalogens, and insufficient carnitines, it can indicate adaptation to lipotoxicity. |
| Sphingomyelins (SM) | SM 34:7;O3<br>SM 35:7<br>SM 49:0;O2<br>SM 65:3 | SM supports membrane structure and microdomains signaling (receptors, ion channels, signaling proteins). Its levels reflect ceramide metabolism. Elevated SM may indicate compensation against ceramide toxicity. The accompanying rise in DAG as CER is |
|  |  | converted to SM can also induce ER stress and promote lipotoxic and pro-inflammatory signaling. |
| Cholestone | Cholest-4-en-3-one,<br>Cholestone | Cholestone are oxidized cholesterol. Elevated level of cholestone is a downstream marker of high oxidative stress, activated macrophages, impaired cholesterol export, unstable high-density lipoprotein (HDL), and sterol-protective system failure. |
| Triacylglycerides (TG) | TG 77:7<br>TG 70:6<br>TG 51:3<br>TG 79:6 | TG is a type of storage lipid carrying fatty acids. However, increased TG level is associated with high TG transport activity, high hepatic very low-density lipoprotein (VLDL) production, and slow TG clearance rate. Insulin resistance, reduced mitochondria beta-oxidation, and impaired fatty acid oxidation (e.g. leading to low carnitines) can be linked with accumulated TG in ceramide-induced lipotoxic condition. |
| Hexosylceramides<br>(HexCer) | HexCer 39:0<br>HexCer 66:4<br>HexCer 31:0 | HexCer are CER with sugar, and acyl-hexosyl-ceramide (AHexCer) are HexCer with a fatty-acyl chain. They are glycosphingolipids that involve structural, signaling, and metabolism roles. Increased HexCer |
|  | AHexcer 75:12 | level suggests active detoxification of CER into glycosphingolipids, |
|  | AHexCer 78:12 | but it signals poor protective response rather than efficient CER |
|  | AHexCer 87:0 | detoxification. |

### Lipidomics in FE-SDMD versus controls

The preliminary metabolomics collection contained 355 identified lipids. As a result, we compiled a curated lipid data set, excluding exogenous, drug-related, artifact-derived, or low-confidence metabolites, ultimately retaining 254 lipids in the final data file. Accordingly, we executed RFECV utilizing random forest on the lipids, ranking the variables by a SHAP summary plot, discarding the least significant features, and cross-validating the accuracy. Consequently, 48 lipids with elevated confidence were selected. The Random Forest model demonstrated satisfactory selection, with an overall accuracy of 88.6%. The training demonstrated a sensitivity of 97.1% and a specificity of 96.6%, whereas the testing samples exhibited a sensitivity of 70% and a specificity of 66.7%.

**Figure 1** displays a heatmap that depicts the hierarchical clustering of the 48 optimal lipids categorized by FE-SDMD and controls. In a significant proportion of FE-SDMD patients, discrete, block-like clusters of elevated lipids are observable throughout the matrix. Nevertheless, a small subset of FE-SDMD patients demonstrates a lipid profile similar to that of the controls.

**Figure 1.**
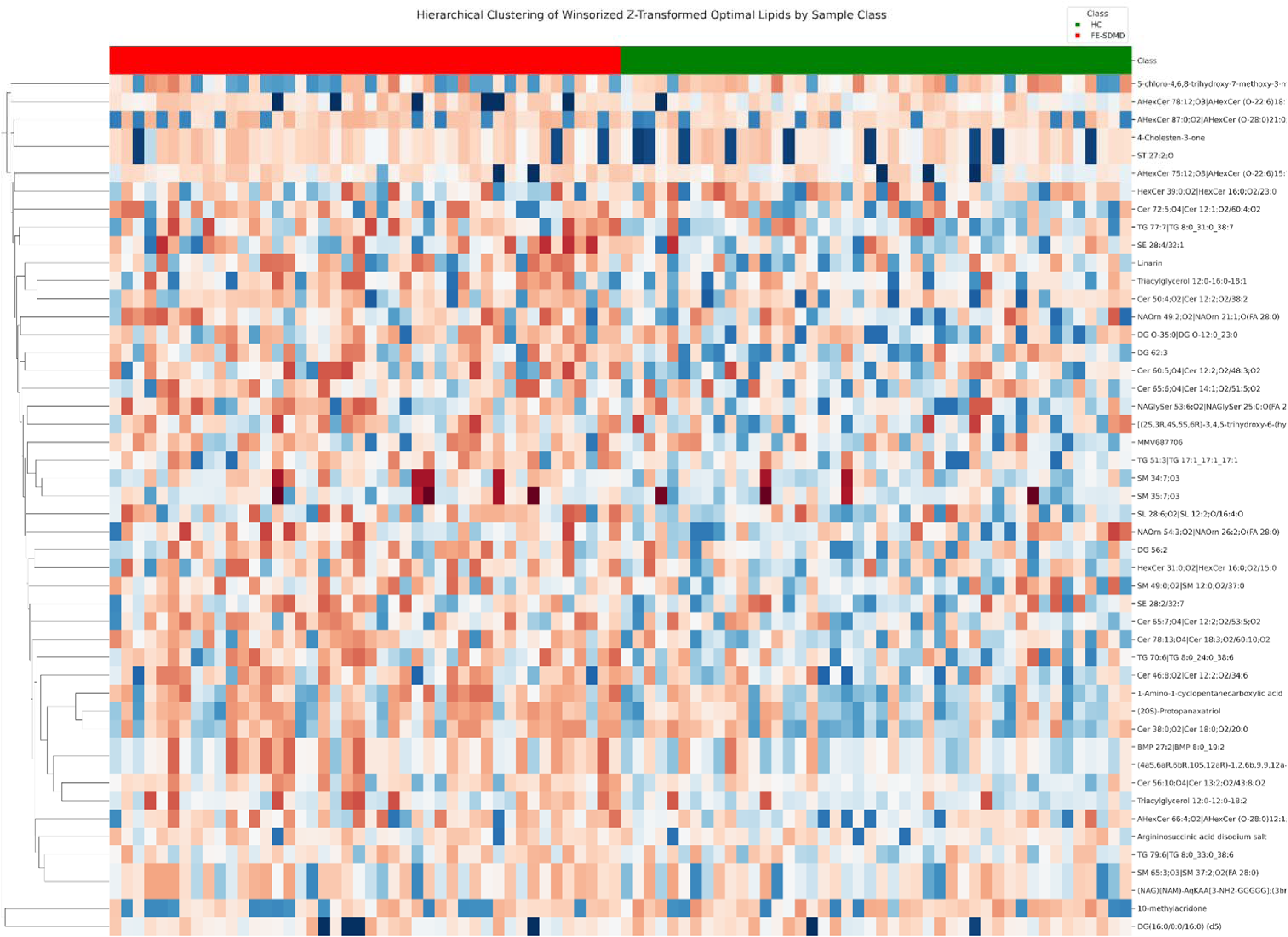
Hierarchical clustering heatmap of z-transformed lipid profiles in FE-SDMD and healthy controls (HC). Rows represent individual lipid species, and columns represent subjects. Red/blue coloration indicates higher/lower relative abundance. Dendrogram derived from hierarchical clustering showing relationships between lipid species. Color coding along the top of the heatmap highlights group-level separation.

To obtain a purer signal for the construction of meaningful lipid modules, we again iteratively eliminated less relevant lipids, as assessed by the Wilcoxon test, PLS-DA, and Random Forest, to create a more parsimonious dataset with an interpretable signature conducive to the development of functional lipid modules. This method diminished the data set to 28 lipids. **Table 1** presents the eight created modules, their respective constituents, and the principal functions of these modules (also see ESF Table 2 for more details). **Table 2** illustrates the differences in lipid module scores between FE-SDMD and the control group. The multivariate GLM indicates significant differences in lipid modules between FE-SDMD and controls (F=5.84, df=8/78, p<0.001, R^2^=0.375). Age, sex and BMI were not significant, but smoking had a significant yet minor influence (F=2.31, df=8/78, p=0.028, R^2^=0.191). DAG, CER, BMP, TG, SM, fatty-acyl amino acids (FAAA), and cholestone significantly showed upregulation in FE-SDMD relative to controls. Hexosylceramides (HexCer) levels in FE-SDMD and control groups were not significantly different. These findings persisted even after applying FDR correction. We also examined possible effects of the drug state of the patients on the 8 module scores but could not detect any effects of use of sertraline (n=15), escitalopram (n=11), other antidepressants (including fluoxetine, bupropion and venlafaxine: n=12), benzodiazepines (n=21), and atypical antipsychotics (n=8). Only two patients were taking mood stabilizers. Therefore, there is no evidence that the drug state of the patients had affected the results.

**Table 2.** Univariate analysis of lipid clusters differences between patients with first-episode simple dysmood disorder (FE-SDMD) and healthy controls (HC)

| **Variables** | **HC**<br>**(n=44)** | **FE-SDMD**<br>**(n=44)** | ***F***<br>**(df=1/85)** | ***p*-value** | **FDR-**<br>**adjusted**<br>***p*-value** | **Partial**<br>**$\eta^2$** |
| --- | --- | --- | --- | --- | --- | --- |
| Diacylglycerides | -0.454 (0.137) | 0.454 (0.137) | 21.72 | <.001 | 0.002 | 0.204 |
| Ceramides | -0.434 (0.138) | 0.434 (0.138) | 19.58 | <.001 | 0.002 | 0.187 |
| BMP 27:2 | -0.416 (0.140) | 0.416 (0.140) | 17.44 | <.001 | 0.002 | 0.17 |
| Triacylglycerides | -0.386 (0.142) | 0.386 (0.142) | 14.55 | <.001 | 0.002 | 0.146 |
| Sphingomyelins | -0.351 (0.144) | 0.351 (0.144) | 11.64 | <.001 | 0.002 | 0.12 |
| Fatty-acyl Amino Acids | -0.253 (0.149) | 0.253 (0.149) | 5.68 | 0.019 | 0.025 | 0.063 |
| Cholestone | -0.226 (0.150) | 0.226 (0.150) | 4.46 | 0.038 | 0.043 | 0.05 |
| HexosylCeramides | -0.053 (0.144) | 0.053 (0.144) | 0.27 | 0.606 | 0.606 | 0.003 |

*Note.* Results are mean and standard errors using ANOVA to compute pairwise comparisons. Variables in between-subject tests are z-scores.
Univariate effects adjusted for Smoking.

**Figure 2** illustrates the outcome of PLS-DA conducted on these 28 lipids. PC1 was the most discriminative component (t1), representing 15.73% of the variation, whilst t2 accounts for 5.032% of the discriminative variance. Figure 2A presents a two-dimensional plot that reveals a different pattern between the two groups, despite a notable overlap between the classes. A notable multivariate distinction was achieved between the two classes (R²Y = 0.38; R²X = 0.16, Q² = 0.28; p < 0.01). Permutation testing (Figure 2B) validated the model, since all permuted models exhibited significantly inferior performance compared to the empirical model, resulting in a negative intercept for Q2 (Q² = −0.3565), thus ruling out chance-driven group separation. The PLS-DA exhibited an accuracy of 79.5%, a sensitivity of 61.4%, and a specificity of 97.7%. The AUC ROC was 0.92 ± 0.058 (Gini index = 0.841, maximum K-S = 0.682). Figure 3 illustrates the VIP scores derived from the aforementioned PLS-DA study. HexCer 39:0, Cer 38:0, BMP 27:2, Cer 56:10, and DG 56:2 were the five most significant lipids influencing the differentiation between FE-SDMD and control groups.

**Figure 2.**
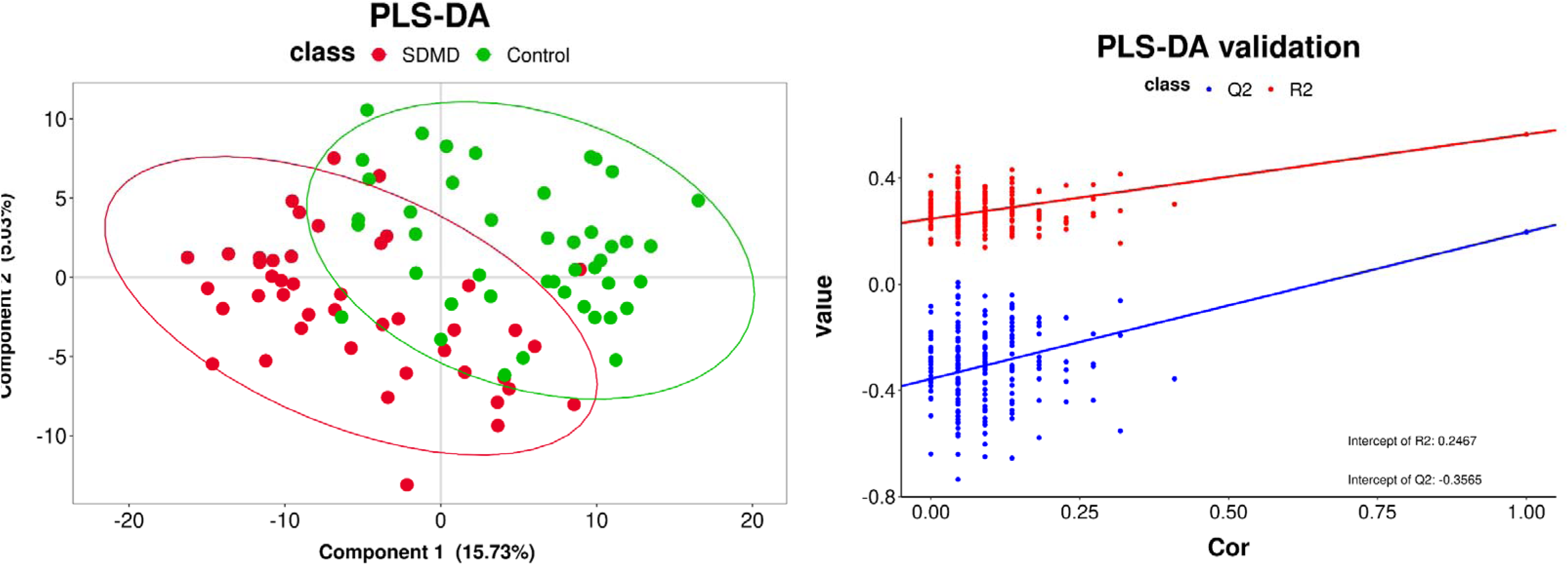
Partial least squares discriminant analysis (PLS-DA) score plot showing separation between FE-SDMD (red) and controls (green) based on the most important lipids. Ellipses indicate 95% confidence intervals for each group, highlighting multivariate lipid differences between FE-SDMD and controls. Right panel: PLS-DA model validation plot showing correlation between predicted and observed values. Red dots and line represent R²Y, and blue dots and line represent Q². Intercepts of R²Y (0.2467) and Q² (–0.3565) demonstrate model robustness and lack of overfitting.

**Figure 3.**
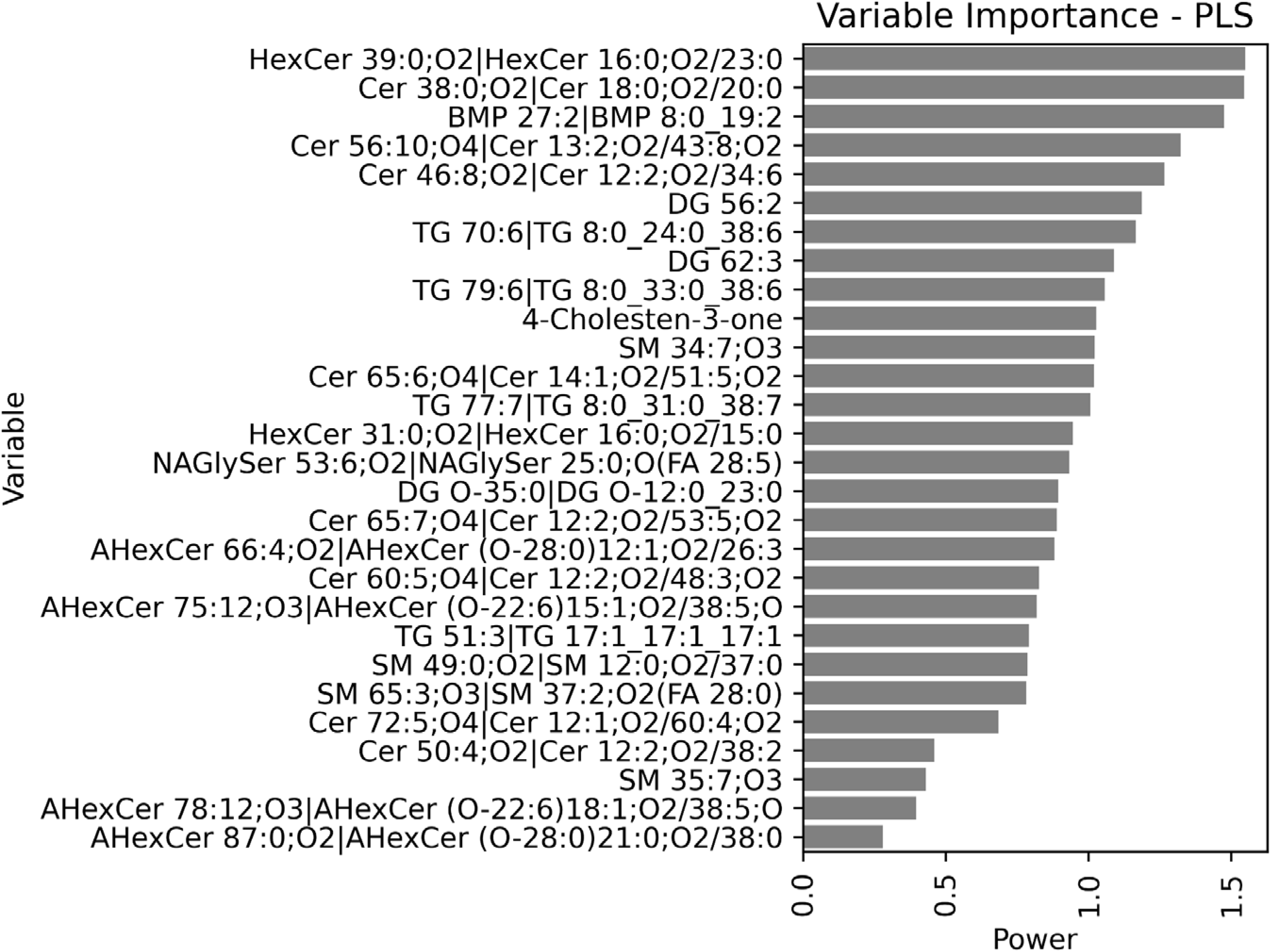
Variable importance in PLS-DA for FE-SDMD versus healthy controls. Bar plot showing the power scores of the top lipid species contributing to class separation in the PLS-DA model. Higher scores indicate greater contribution of the lipid to distinguishing FE-SDMD (red) from healthy controls (green). Key lipids include ceramides (CER), hexosylceramides (HexCer), bis-monoacylglycerol phosphate (BMP), diacylglycerides (DG), triacylglycerides (TG), sphingomyelins (SM), and N-acylglycine-serine (NAGlySer).

### ACEs, Lipidomics and phenome scores

**Figure 4** displays a correlation heatmap that depicts Pearson’s correlations among the 8 modules and clinical data, including psychological stresses and phenome scores. ACEs were linked to CER, SM, and BMP. NLEs were correlated with CER, DAG, cholestone, BMP, and HexCer. The primary NLE predicting cholestone was academic NLEs, accounting for 17.6% of the variation (F=18.20, df=1/85, p<0.001). Total NLEs accounted for 12.8% of variance in CER levels (F=12.53, df=1.85, p<0.001). The product term ACEs × NLEs was linked to all modules, with the exception of TG, and was particularly correlated with CER levels. The four phenome domains were associated with all eight modules, except for pure BDI and triacylglycerides, physiosomatic and cholestone and suicidal behaviors and HexCer.

**Figure 4.**
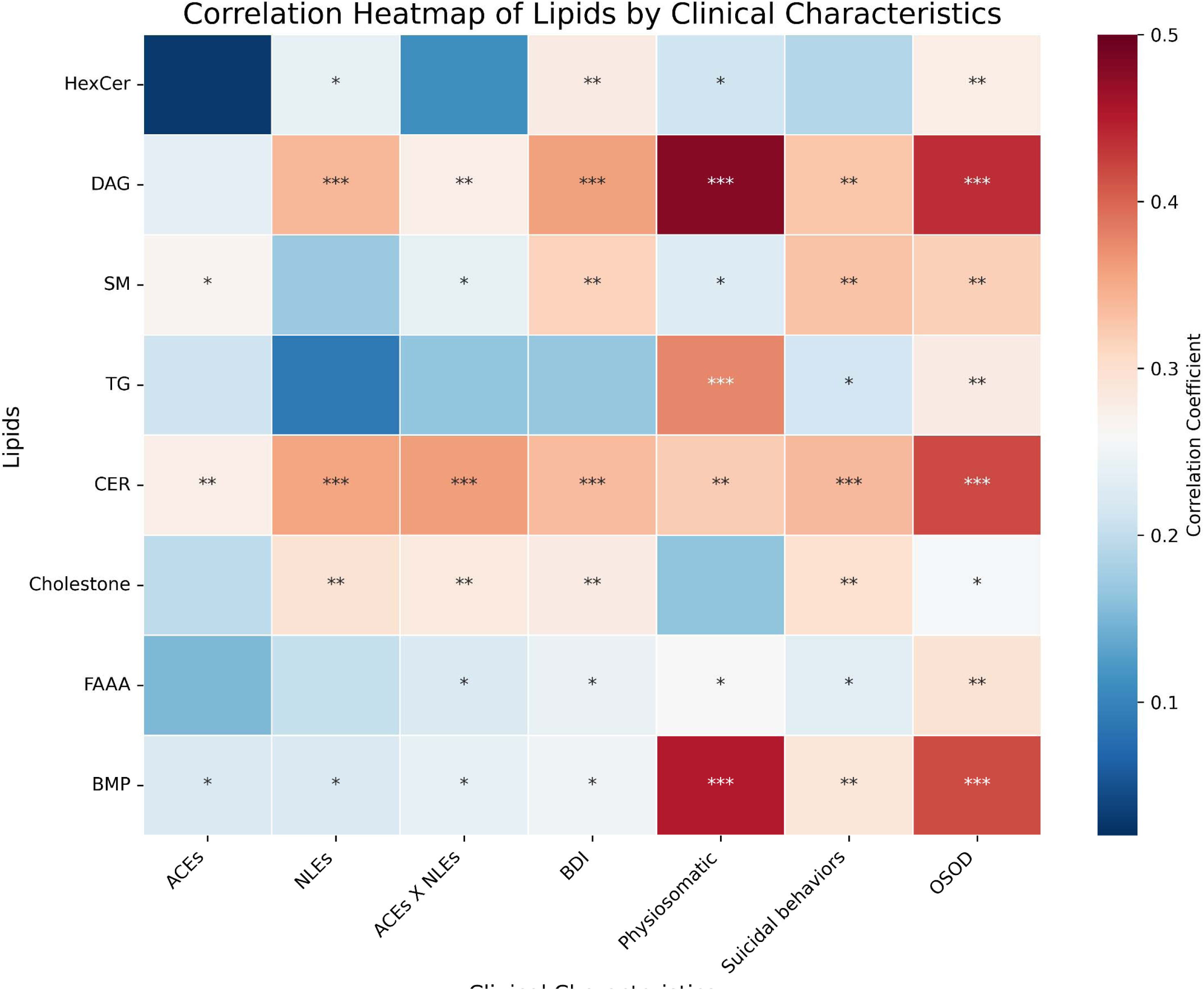
Correlation heatmap of lipid modules with clinical characteristics in FE-SDMD. Heatmap displaying Pearson correlation coefficients between key lipid species (HexCer, CER, DAG, SM, TG, FAAA, BMP, cholestone) and clinical characteristics, including adverse childhood experiences (ACEs), negative life events (NLEs), ACEs × NLEs interaction, pure BDI score, physiosomatic symptoms, suicidal behaviors, and OSOD. Red/blue coloration represents correlations, and asterisks indicate significance levels (*p < 0.05; **p < 0.01; ***p < 0.001). This figure highlights the association between lipotoxic and compensatory lipid modules and clinical symptom domains, emphasizing the influence of stressors on lipid dysregulation in FE-SDMD.

To investigate the impact of lipidomics on phenome scores, we conducted PLS regression, using the phenome scores (OSOD, physiosomatic symptom domain, suicidal behaviors, and pure BDI scores) as dependent variables and the eight functional lipidomic modules as independent variables. Figure 5A presents the outcomes of those PLS regression analyses. Figure 5A illustrates that a notable multivariate prediction was achieved when predicting OSOD (R²Y = 0.35; R²X = 0.34; Q² = 0.30; p < 0.01). The three primary markers, in descending order, were DAG, CER, and BMP. Figure 5B indicates that 24.8% of the variance in suicidal behaviors correlates with the component derived from the eight lipidomics profiles (R^2^X=0.34, Q² = 0.16; p < 0.01). The three primary markers, listed in descending order, were cholestone, CER, and SM. A significant portion of the variance in the pure BDI score (Figure 5C) was elucidated by the first PLS-regression component (R²Y = 0.26; R²X = 0.34; Q² = 0.18; p < 0.01). The DAG module was the most significant, followed by CER and cholestone. A significant portion of the variance in the physiosomatic symptom domain (Figure 5D) was elucidated by the first component derived from the modules (R²Y = 0.32; R²X = 0.34; Q² =0.28; p < 0.01), with DAG identified as the predominant factor, followed by BMP and CER.

**Figure 5.**
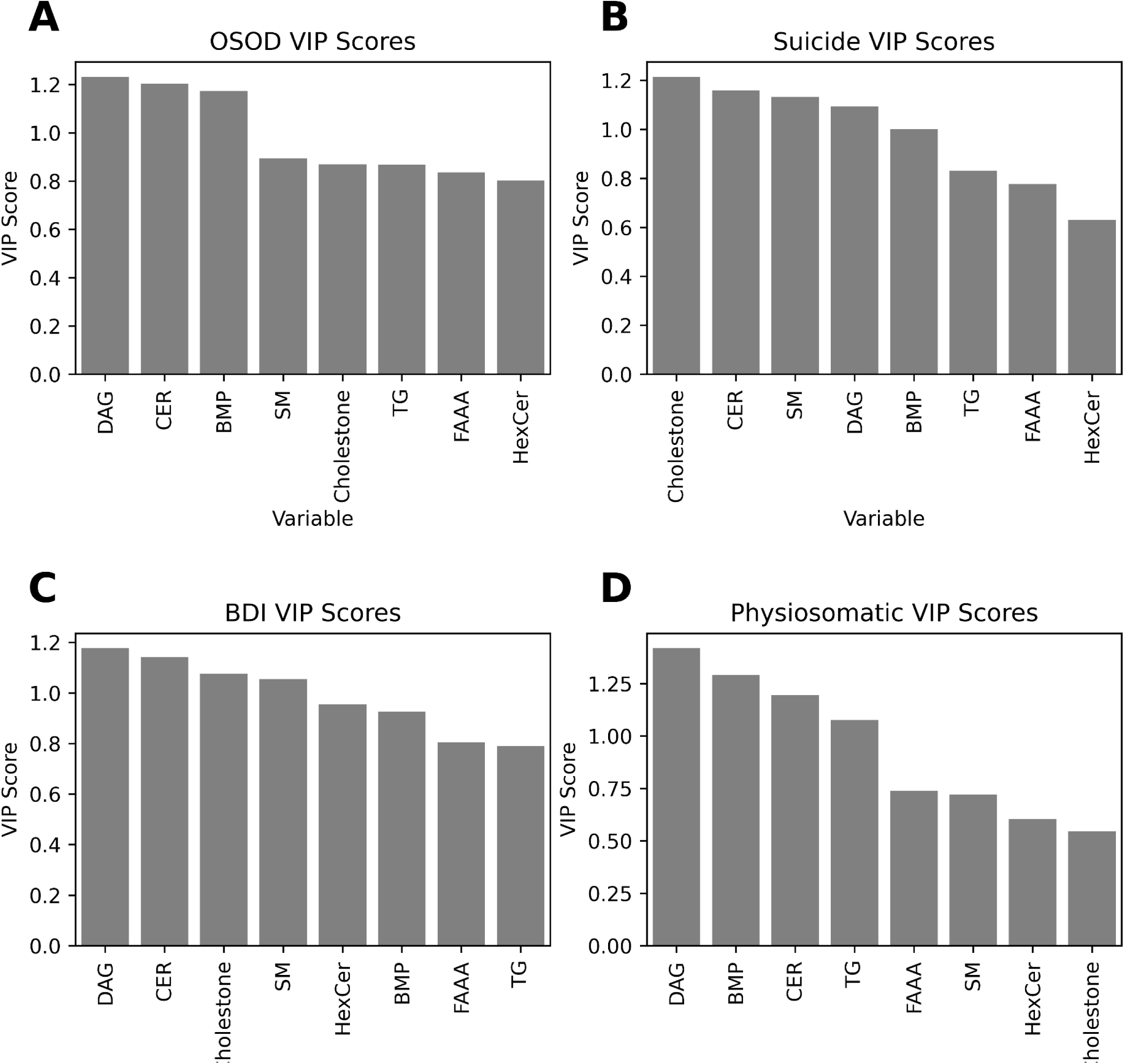
Variable importance in projection (VIP) scores in PLS regression for clinical phenotypes in FE-SDMD. Bar plots showing VIP scores for key lipid species predicting different key clinical phenome outcomes, i.e. OSOD: overall severity of illness, suicidal behaviors, pure BDI scores, and physiosomatic symptoms. Key contributing lipids include ceramides (CER), hexosylceramides (HexCer), diacylglycerides (DAG), triacylglycerides (TG), sphingomyelins (SM), bis-monoacylglycerol phosphate (BMP), cholestone, and N-acylglycine-serine (FAAA). Higher VIP scores indicate greater predictive contribution of the lipid to the specific clinical phenotype.

### Lipidomics, NIMETOX biomarkers and phenome scores

First, we examined the associations between ONS and LCAT indices and the 8 modules. No significant correlations could be established. Consequently, we examined the effects of the 8 lipid modules, and ONS and LCAT variables on the phenome domains of FE-SDMD. Toward this end, we employed manual and forward-stepwise multiple regression analysis with an entry probability of ≤ 0.05 and a deletion probability of ≥ 0.1. **Table 3** presents the outcomes of the models wherein clinical outcomes serve as dependent variables, with lipid domains, additional biomarkers (ONS index; LCAT index), and clinical-demographic variables (age, sex, smoking) as covariates. DAG, BMP, cholestone, ONS Index, and LCAT accounted for 46.4% of the variance in the OSOD score. DAG, ONS Index, BMP, and LCAT elucidated 42.4% of the variance in physiosomatic symptoms. DAG, ONS Index, SM, cholestone, LCAT, and HexCer concentrations accounted for 37.9% of the variance in the pure BDI score. CER, LCAT, cholestone, and SM accounted for 30.0% of the variance in PC lifetime suicidal behaviors. BMP, atherogenicity, DAG, and smoking status accounted for 32.0% of the variance in the ISI score. CER and LCAT accounted for 19.5% of the variance in the total state anxiety score.

**Table 3.**
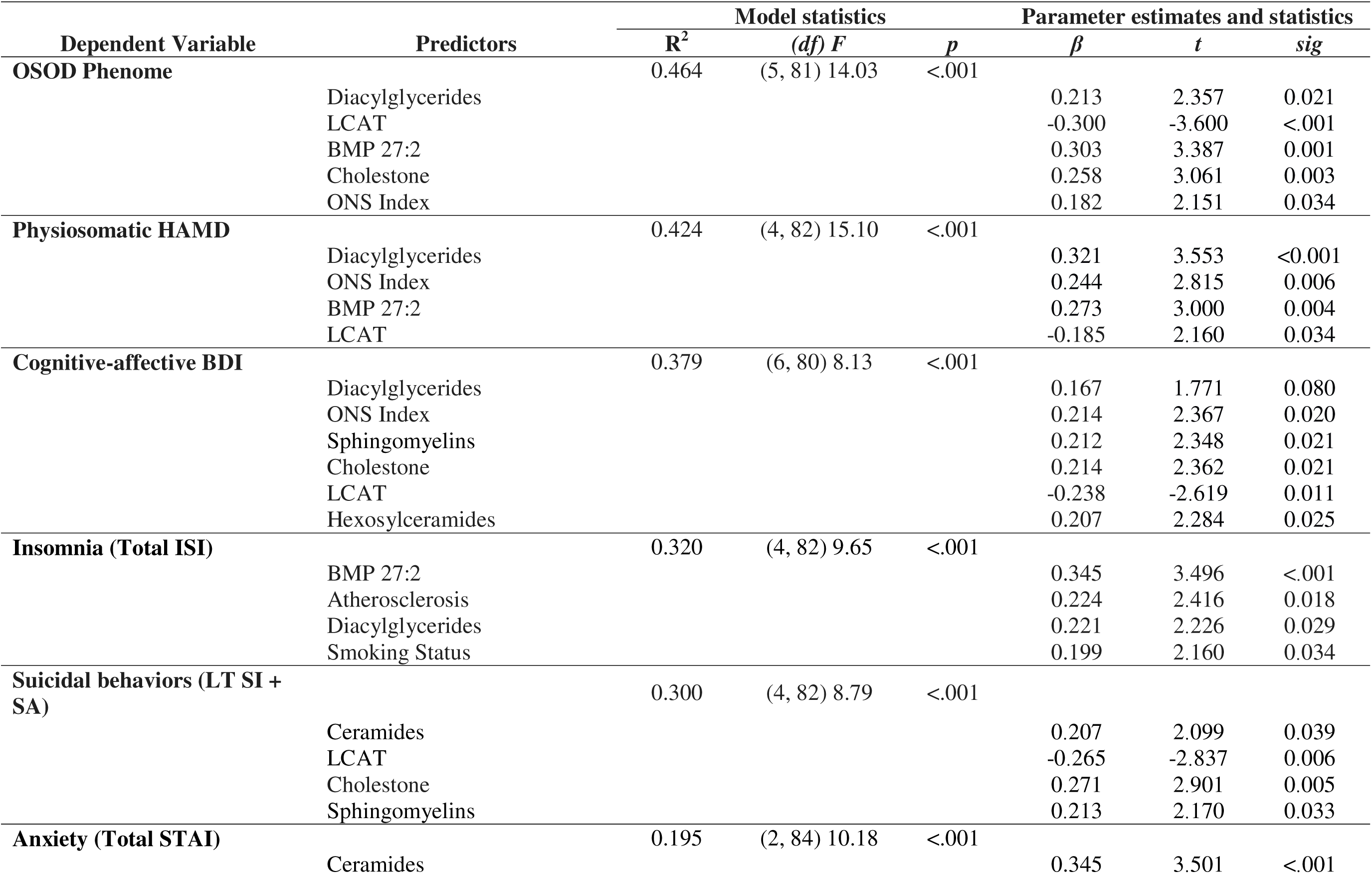
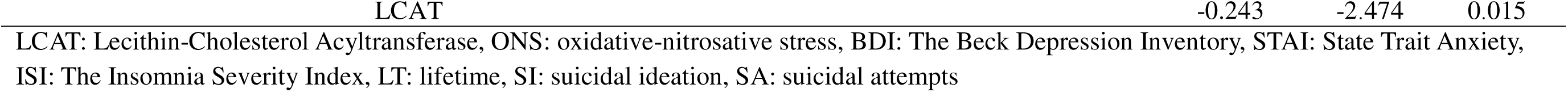
Multiple regression models predicting clinical phenotypes using lipid modules and other biomarkers as explanatory variables.

### Personalized approach

**Figure 6** illustrates the score contribution profiles for four individuals: one healthy control (Figure 6A) and three patients with FE-SDMD (Figures 6B, 6C, and 6D). These data underscore the contributions of the lipidomic modules to the differentiation between the SDMD and healthy control classes. The latter relies on PLS-DA shown above. The module scores were all below zero in the healthy control group. Significant heterogeneity was seen between the three MDD patient profiles. Thus, in certain SDMD patients, CER predominates, whereas in others, there is an elevation in DAG and FAAA.

**Figure 6.**
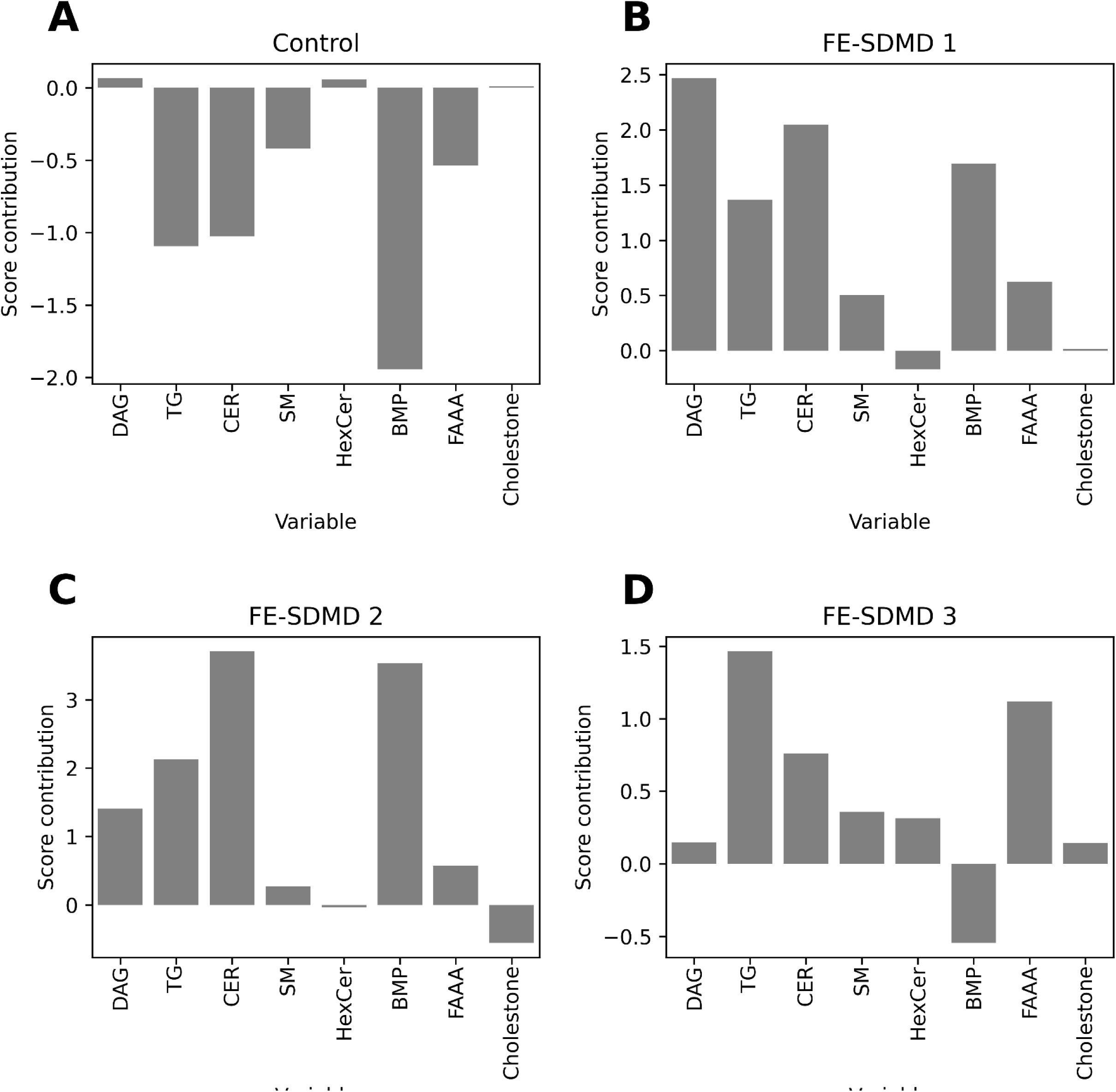
Personalized VIP score contributions for controls and individual FE-SDMD patients. The bar plots show the contributions of key lipid species to PLS-DA scores on a per-subject basis. (A) Control participant, illustrating baseline lipid contributions. (B–D) Three representative FE-SDMD patients, highlighting individual patterns of lipotoxic and compensatory lipid modules. These plots demonstrate inter-individual variability in lipid contributions, reflecting personalized lipid dysregulation profiles in FE-SDMD. CER: ceramides (CER); HexCer, hexosylceramides; DAG: diacylglycerides; TG: triacylglycerides, SM: sphingomyelins, BMP: bis-monoacylglycerol phosphate; FAAA: fatty-acid amino-acids

## Discussion

### Key functional Lipid modules in FE-SDMD

Through lipidomics analysis, we found seven lipid modules that were significantly upregulated in FE-SDMD, i.e., CER, DAG, TG, SM, BMP, cholestone, and FAAA. Earlier lipidomic studies in MDD reported reduced acylcarnitines and SM, coupled with increased CER and oxidized fatty acids (He et al., 2025; Liu et al., 2016; Miao et al., 2023; F. Wang et al., 2022; Zhang et al., 2022). A recent review summarized that MDD patients showed alterations in CER, SM, PC, LPC, and LysoPE (Zorkina et al., 2024). However, previous findings are sometimes conflicting in terms of the direction of lysophospholipids (e.g., LPC, LPE, and lysophosphatidylinositol; LPI) and phospholipids (e.g., PC, PE, and phosphatidylinositol; PI). Some studies reported overall upregulation (Liu et al., 2016; Miao et al., 2023), while others reported downregulation (Kim et al., 2018; Okamoto et al., 2025; F. Wang et al., 2022; J. W. Zhang et al., 2026). Results regarding glycerolipids levels were also conflicting (He et al., 2025; Kim et al., 2018; Liu et al., 2016).

One major issue is that semi-quantitative LC-MS lipidomics data may display semi-compositional characteristics due to ion competition, ion suppression, normalization procedures, and relative abundance measurements. Consequently, apparent increases in some lipid species or classes may induce relative decreases in others, particularly when (semi-)compositional effects are not addressed using approaches such as CLR (or alternatively isometric log-ratio or additive log-ratio) transformations. Thus, these transformations are useful not because lipidomics is fully compositional, but because the data can behave partially compositionally in practice. Other factors may contribute to the discrepant results in MDD, including heterogeneity in MDD, medication effects, MetS, ROI-dependent neurobiology, exclusion of medical comorbid disorders, lipidomics methods such as positive versus negative ion mode, statistical approaches, diet, ethnicity, and lifestyle.

In this context, it is instructive to compare two well-controlled studies, the present study and Y. Zhang et al. (2026). Both applied CLR transformation, similar exclusion criteria, and controlled for metabolic confounders such as MetS (no participants with MetS were included in the current study). Notably, despite differences in clinical severity, both studies observed elevations in CER, DAG, TG, and SM. The effect sizes, however, were more pronounced in Y. Zhang et al. (2026), which included severely depressed inpatients, many with increased recurrence of illness (ROI). These observations suggest that lipid dysregulation may worsen with greater clinical severity and higher ROI, consistent with prior evidence linking ROI to impaired RCT and increased atherogenicity (Maes, Niu, Maes, et al., 2026).

### Lipidomics and the depression phenome

Importantly, the lipid modules were consistently related to the key clinical domains of FE-SDMD, including OSOD, physiosomatic symptoms, a restricted cognitive-affective score (pure BDI), suicidal behaviors, insomnia (ISI), and state anxiety (STAI score). Our findings indicate that lipid aberrations strongly contribute to these domains beyond and above the effects of ONS and LCAT. The ONS environment and lower LCAT/RCT likely drives the accelerated production and accumulation of toxic lipid species, creating a self-reinforcing cycle of ROS signaling, membrane dysfunction, cellular stress, and eventually immune-inflammatory responses (Amarjeet et al., 2023; Brinholi et al., 2024; Horton et al., 1987; Li et al., 2025; Zorkina et al., 2024).

It should be emphasized, however, that the current FE-SDMD sample showed no overt signs of systemic activation of the IRS; instead, there was a reduction in CIRS activity (Maes, Jirakran, et al., 2024). This suggests that early-stage SDMD may be characterized more by an impaired ability to regulate immune responses than by an active IRS response. In this context, decreased LCAT activity and impaired RCT, together with the consequent accumulation of lipotoxic species such as CER, DAG, SM, and oxidized sterols, may create a pro-oxidative and lipotoxic environment. These alterations may gradually sensitize the IRS, lowering the threshold for its activation. Over time, particularly in patients with increased ROI, the combined effects of chronic oxidative–nitrosative stress, lipotoxicity, and inadequate CIRS responses may drive sustained IRS activation (Maes, Almulla, You, et al., 2025). Thus, lipid dysregulation and oxidative stress may act as early mechanistic contributors that set the stage for later IRS activation in the course of depression.

### Psychological stress and lipidomics

ACEs were positively associated with CER, SM, DAG, cholestone, and BMP, extending previous findings that ACEs are associated with RCT, increased atherogenicity, and lipotoxicity (Maes, Jirakran, et al., 2024). ACEs are also known to predispose individuals to future metabolic and mental health problems through upregulated inflammatory markers—such as interleukin-6, C-reactive protein, fibrinogen, and intercellular adhesion molecule-1—as well as stress-related hormones, including glucocorticoids and catecholamines (Alley et al., 2025; Liu et al., 2026). These inflammatory and stress pathways have been linked to CER and DAG metabolism (de Mello et al., 2009; Peckett et al., 2011).

CER, DAG, cholestone, BMP, and HexCer were also positively associated with NLEs, with academic stress specifically correlating with cholestone, reflecting oxidation of free cholesterol and systemic oxidative stress. Moreover, the interaction between ACEs × NLEs amplified associations with cholestone, CER, DAG, BMP, SM, and FAAA. These results indicate that psychological stressors in FE-SDMD not only alter oxidative-nitrosative stress markers, LCAT activity, and atherogenicity (Brinholi et al., 2024; Maes, Almulla, You, et al., 2025; Maes, Vasupanrajit, et al., 2024) but also profoundly reshape key lipid profiles, highlighting the central role of stress-induced lipotoxicity in early depression.

### Mechanistic explanations

CER accumulation may exert neurotoxic effects via activation of NADPH oxidase, mitochondrial dysfunction, ROS production, and apoptosis (Amarjeet et al., 2023; Barth et al., 2012). DAG and TG act as lipid second messengers and accumulate during CER metabolism. DAG activates Protein Kinase C, impairing insulin receptor signaling and glucose transporter function, while DAG and saturated fatty acids stimulate proinflammatory cytokines via c-Jun N-terminal kinase/IκB kinase β/NF-κB pathways (Eichmann & Lass, 2015; Ge et al., 2025). Oxidized sterols, such as cholestone, increase under ONS, disrupt lipid rafts, impair transforming growth factor-β receptor signaling, and enhance ER stress and ROS production, exacerbating apoptosis and inflammation (Chen et al., 2017; Vejux & Lizard, 2009). BMP, a lysosomal lipid mediator, rises in response to toxic lipid overload, activating hydrolases and reflecting lysosomal stress (Medoh & Abu-Remaileh, 2024). Therefore, BMP upregulation in FE-SDMD, especially in patients with insomnia, physiosomatic symptoms, and high OSOD, parallels elevations in CER, SM, and cholestone, possibly indicating lysosomal overload.

Our findings may also suggest that depressive pathology may entail disrupted compensatory mechanisms aimed at counteracting CER-driven lipotoxicity (Amarjeet et al., 2023; Barth et al., 2012; Bernal-Vega et al., 2023). CER is partially metabolized to SM via sphingomyelin synthase, a compensatory mechanism to neutralize the toxic effects of CER. While this reduces CER toxicity, excessive SM can rigidify membranes and disrupt signaling (Arsenault et al., 2021; Kalinichenko et al., 2026). Glycosylation of CER to HexCer provides another detoxification route; however, increases in HexCer may have apoptotic, cytotoxic, neuroinflammatory potential and disrupt membrane and signaling (Borodzicz et al., 2015; Düsing et al., 2023; Ishibashi et al., 2013). TG accumulation buffers DAG toxicity but contributes to neuroinflammation and correlates with depressive symptoms (Bot et al., 2020; Luan et al., 2025). FAAA (e.g., NAGlySer) serve as endocannabinoid-like modulators of inflammation. Although not selected in stepwise models, FAAA levels correlated with OSOD, depressive severity, physiosomatic symptoms, and suicidal behaviors. FAAA elevations may act as a compensatory response to overall lipotoxic stress (Anderson & Merkler, 2017; Battista et al., 2019). Nevertheless, the mild increase in FAAA observed here may interfere with immune-inflammatory pathways and pain (Burstein, 2018).

Collectively, the accumulation of CER, DAG, and oxidized sterols occurs alongside reduced LCAT activity, impairing HDL maturation and RCT. This amplifies ONS, disrupts membrane homeostasis, and promotes inflammatory signaling. These lipid alterations operate within an ONS environment, accelerating toxic lipid production, creating a self-reinforcing cycle of NIMETOX dysregulation contributing to the pathophysiology of FE-SDMD (Amarjeet et al., 2023; Brinholi et al., 2024; Horton et al., 1987; Li et al., 2025; Maes et al., 1994; Maes, Niu, Maes, et al., 2026; Zorkina et al., 2024).

### Limitations

The current study has some limitations. First, recruitment was limited to university students, which may restrict generalizability to other age groups and/or populations. Second, by focusing on first-episode depression, we minimized confounding from long-term antidepressant use but the findings may not reflect metabolic or lipolytic alterations observed in chronic or recurrent MDD. Third, its cross-sectional design does not allow it to firmly establish causal inferences and does not reveal whether lipid changes precede or follow with depressive symptoms. The case–control design limits precise mechanistic interpretation of relationships among lipid modules, oxidative–nitrosative stress markers, atherosclerosis indicators, and depressive phenotypes. Longitudinal studies are needed to clarify the temporal dynamics of lipid dysregulation in MDD. Fourth, serum lipid profiling was performed in positive ion mode, which may underrepresent acidic phospholipids and other negatively ionizing species. However, previous studies demonstrate that positive ion mode reliably detects lipids relevant to MDD (Kim et al., 2018; Lee et al., 2013).

### Conclusion

FE-SDMD is characterized by early and distinct lipid dysregulations, including elevated CER, DAG, TG, SM, cholestone, BMP, and HexCer. Lipotoxic CER and DAG promote neurotoxic, metabolic, and inflammatory processes, whereas SM, HexCer, and FAAA may reflect compensatory detoxification mechanisms. Psychological stressors, including ACEs and NLEs, are positively associated with these lipid modules, linking cumulative psychological stressors to early lipid dysregulation. Notably, interactions between ACEs and NLEs further amplify these lipid alterations, suggesting that combined early-life and recent stress exacerbate lipotoxicity. Lipid dysregulation, together with ONS and reduced LCAT activity, correlates with core clinical phenotypes, including depressive severity, physiosomatic symptoms, suicidal behaviors, anxiety and insomnia. These findings support the NIMETOX framework, highlighting lipid-mediated mechanisms as early contributors to neuroimmune–metabolic disturbances in MDD. Importantly, early lipid abnormalities may serve as biomarkers for disease onset and prognosis, offering potential targets for precision psychiatry interventions.

Previously, Maes, Almulla, Stoyanov, et al. (2025) reviewed evidence indicating that combined interventions with PPAR agonists, statins, omega-3 fatty acids, and LCAT activators may offer therapeutic benefit in MDD by directly targeting lipid dysregulation. PPAR agonists can improve fatty acid oxidation and reduce ceramide and DAG accumulation, statins limit oxidized sterol buildup and systemic inflammation, omega-3 fatty acids support membrane integrity and modulate lipotoxic pathways, and LCAT activators improve HDL maturation and RCT. Together, these approaches could mitigate lipotoxicity, reduce ONS, and restore neuroimmune–metabolic homeostasis.

## CRediT authorship contribution statement

**Visuthsiri Sirivatanapa:** Conceptualization, Data Curation, Formal Analysis, Funding Acquisition, Investigation, Project Administration, Software, Visualization, Writing – original draft. **Pannipa Janta:** Data Curation, Methodology, Resources, Validation. **Asara Vasupanrajit:** Project Administration, Resources, Investigation. **Chavit Tunvirachaisakul:** Supervision, Resources, Writing – Review & Editing. **Sira Sriswasdi:** Software, Supervision. **Rossarin Tansawat:** Resources, Supervision. **Andre F Carvalho**: Review, Editing, Validation. **Yingqian Zhang:** Software, Visualization. **Michael Maes:** Conceptualization, Data Curation, Formal Analysis, Methodology, Project Administration, Supervision, Validation, Visualization, Writing – original draft, Writing – Review & Editing.

## Ethics approval and consent to participate

The research project (#445/63 and #054/68) was approved by the Institutional Review Board of Chulalongkorn University’s institutional ethics board, Bangkok, Thailand, which follows the International Guideline for Human Research protection as required by the Declaration of Helsinki, The Belmont Report, CIOMS Guideline and International Conference on Harmonization in Good Clinical Practice (ICH-GCP). All participants gave written informed consent prior to participation in the study (#445/63).

## Supporting information

Electronic Supplementary File

## Data Availability

Datasets analyzed in the present study will be made available upon reasonable request to the corresponding authors after all the data have been fully processed

