## Supplementary material for "Psychological Stress-Associated Ceramide and Diacylglyceride Lipotoxicity as Contributors to First Episode Depression Pathophysiology: A neuroimmune-Metabolic-Oxidative Stress (NIMETOX) Perspective": Electronic Supplementary File

**ESF Table 1**. Participant’s demographic data, clinical characteristics, and biomarkers

| **Variables** | **Controls (n=44)**  **Mean (SD)** | **FE-SDMD (n=44)**  **Mean (SD)** | ***p*** |
| --- | --- | --- | --- |
| Age (years) | 23.477 (3.181) | 21.933 (3.298) | 0.028 |
| Sex,^a^ n (%) |  |  |  |
| Female | 37 (84.1%) | 38 (86.4%) | 0.764^a^ |
| Male | 7 (15.9%) | 6 (13.6%) |  |
| Year of education | 16.568 (1.485) | 15.523 (2.799) | 0.031 |
| BMI (kg/m^2^) | 22.331 (3.574) | 21.705 (4.899) | 0.495 |
| Current smoking status, ^a^ n (%) |  |  |  |
| No | 43 (97.7%) | 39 (88.6%) | 0.091 ^a^ |
| Yes | 1 (2.3%) | 5 (11.4%) |  |
| Metabolic syndrome, ^a^ n (%) |  |  |  |
| No | 44 (100.0%) | 44 (100.0%) | 1 |
| HAM-D Physio Somatic Symptoms | 0.636 (0.685) | 3.023 (1.355) | <0.001 |
| BDI-II | 6.773 (6.491) | 24.136 (12.233) | <0.001 |
| ISI | 5.591 (4.304) | 12.341 (6.416) | <0.001 |
| Suicidal behavior score ^b^ | -0.932 (0.197) | 0.505 (0.921) | <0.001 |
| ONS index ^b^ | -1.179 (1.714) | 0.795 (2.076) | <0.001 |
| HDL Cholesterol | 60.886 (11.018) | 62.500 (12.446) | 0.521 |
| ApoA | 139.250 (25.770) | 142.318 (26.386) | 0.583 |
| LCAT | 80.728 (3.421) | 78.148 (2.284) | <0.001 |
| RCT | 0.332 (1.984) | -0.067 (2.267) | 0.386 |

T-test and ^a^ Chi-square were used to compute group differences. Data shown in mean and SD. ^b^ mean and SE of z-score. FE-SDMD: first episode simple dysmood disorder; BMI: Body mass index; HAM-D: Hamilton Depression Rating Scale (somatic anxiety + gastro-intestinal somatic + general somatic); BDI-II: The Beck Depression Inventory; ISI: The Insomnia Severity Index; ONS: Oxidative and nitrosative stress; HDL cholesterol: High density lipoprotein cholesterol; ApoA: Apolipoprotein-A; LCAT: Lecithin cholesterol acyl transferase; RCT: Reverse cholesterol transport.

**ESF Table 2**. Construction of eight functional lipid modules

| **Modules** | **Members (synonym)** | **Functions** |
| --- | --- | --- |
| Diacylglycerides (DG) | DG 56:2  DG 62:3  DG O-35 | DG is a potent signaling lipid because it can activate protein kinase C (PKC), which is a central switch to insulin resistance, endoplasmic reticulum (ER) stress, and inflammation. DG is precursor to TG and phospholipid resynthesis. DG is a harmful lipid signaling molecule that disrupts lipid buffering and can act together with ceramides to exacerbate neuroimmune and metabolic stress. |
| Ceramides (CER) | CER 38:0  CER 50:4  CER 56:10  CER 65:6  CER 46:8  CER 72:5  CER 60:5  CER 65:7 | CER function as membrane lipid. At increased level, ceramides are lipotoxic molecules that rigidify membranes, disrupt insulin receptor signaling, permeabilize mitochondria through ceramide channels formation, and block beta-oxidation. Excess ceramides drive cells from adaptive lipid remodeling into inflammatory, apoptotic, and metabolically dysfunctional states. Ceramides can be converted to sphingomyelin and vice versa. |
| Bismonoacylglycerol phosphates (glycerophospholipids; BMP) | BMP 27:2 | BMP is abundant in lysosomal membrane’s intraluminal vesicles, where it supports the sorting and breakdown of lipids, including sphingolipids and cholesterol. BMP activates lysosomal hydrolases, necessary for sphingolipid breakdown. Accordingly, BMP can be viewed as a functional marker of the lysosome’s capacity to process lipids. BMP contributes to the regulation of ceramide metabolism, sphingolipid turnover, and lysosomal cholesterol efflux. Because of this, elevated BMP levels may reflect an increased influx of these lipids into lysosomes. They may also indicate stress on lysosomal degradative pathways, along with compensatory expansion or remodeling of lysosomal membranes. A rise in BMP together with increases in ceramides or hexosyl ceramides, diacylglycerols, and long-chain triacylglycerols can point to chronic lipotoxic stress, increased dependence on lysosomal clearance as a backup pathway, suboptimal resolution of lipid overload, and loss of normal lipid homeostasis. |
| Fatty-acyl amino-acid conjugates (FAAA) | NaGlySer 53:6 | FAAAs like N-acyl glycine-serine conjugate (NaGlySer) are signaling lipids associated with anti-inflammation and detoxification. In response to dysfunctional beta-oxidation, FAAA often increases to reduce excess polyunsaturated fatty acids influx. When co-occur with increased ceramides, decrease plasmalogens, and insufficient carnitines, it can indicate adaptation to lipotoxicity. |
| Sphingomyelins (SM) | SM 34:7;O3  SM 35:7 SM 49:0;O2  SM 65:3 | SM supports membrane structure and organization of signaling microdomains such as receptors, ion channels, and signaling proteins. SM levels indicate how ceramides are processed. SM converts to ceramides via sphingomyelinase, consuming phosphatidylcholine. Ceramide converts to SM via sphingomyelin synthase, producing DG as a byproduct. Accordingly, elevated SM levels may reflect compensatory efforts to counteract rapid ceramide toxicity. The resulting membrane rigidification, driven by more stable structure of SM and the enlargement of cholesterol-enriched lipid rafts, leads to decreased membrane fluidity, disrupted lipid signaling, and dysfunction of synaptic and mitochondrial membranes. In addition, the associated rise in DG as a conversion byproduct can trigger ER stress alongside lipotoxic and pro-inflammatory signaling. |
| Cholestone (Oxidized sterols) | Cholest-4-en-3-one, Cholestone | Cholestone are cholesterol that has been oxidized. Elevated level of oxidized sterols is a downstream marker of high oxidative stress, activated macrophages, impaired cholesterol export, unstable high-density lipoprotein (HDL), and sterol-protective system failure. Although of less importance, oxidized sterols have several biological implications. First, they can infiltrate the cell membrane to interfere with fluidity, raft organization, and signaling. Second, they may disable HDL, increase LDL oxidization susceptibility, and reduce cholesterol efflux. Third, once entering the mitochondria membrane, they can increase reactive oxygen species, reduce electron transport chain efficiency, and produce mitochondrial stress. Fourth, they may signal cell death by inducing ER stress, making cell more vulnerable to lipotoxicity. |
| Triacylglycerides (TG) | TG 77:7 TG 70:6 TG 51:3  TG 79:6 | TG is known as storage lipid carrying fatty acids. However, increased TG level is associated with high TG transport activity, high hepatic very low-density lipoprotein (VLDL) production, and slow TG clearance rate. Insulin resistance, reduced mitochondria beta-oxidation, and impaired fatty acid oxidation (e.g. leading to low carnitines) can be linked with accumulated TG in ceramide-induced lipotoxic condition. |
| Hexosylceramides (HexCer) | HexCer 39:0  HexCer 66:4  HexCer 31:0  AHexcer 75:12  AHexCer 78:12  AHexCer 87:0 | HexCer are ceramides with sugar, and acyl-hexosyl-ceramide (AHexCer) are HexCer with a fatty-acyl chain. They are glycosphingolipids that involve structural, signaling, and metabolism roles. Increased HexCer level suggests active detoxification of ceramides into glycosphingolipids, but it signals poor protective response rather than efficient ceramide detoxification. |
